# Disease progression and clinical outcomes in latent osteoarthritis phenotypes: Data from the Osteoarthritis Initiative

**DOI:** 10.1101/2023.12.14.23299525

**Authors:** Zeyu Huang, Mary A. Bucklin, Weihua Guo, John T. Martin

**Affiliations:** Department of Orthopaedic Surgery, Orthopaedic Research Institute, West China Hospital, West China Medical School, Sichuan University, Chengdu, Sichuan Province, People’s Republic of China; Department of Orthopedic Surgery, Rush University, Chicago, Illinois, USA; Department of Immuno-oncology, City of Hope, National Medical Center, Duarte, California, USA

## Abstract

The prevalence of knee osteoarthritis (OA) is widespread and the heterogeneous patient factors and clinical symptoms in OA patients impede developing personalized treatments for OA patients. In this study, we used unsupervised and supervised machine learning to organize the heterogeneity in knee OA patients and predict disease progression in individuals from the Osteoarthritis Initiative (OAI) dataset. We identified four distinct knee OA phenotypes using unsupervised learning that were defined by nutrition, disability, stiffness, and pain (knee and back) and were strongly related to disease fate. Interestingly, the absence of supplemental vitamins from an individual’s diet was protective from disease progression. Moreover, we established a phenotyping tool and prognostic model from 5 variables (WOMAC disability score of the right knee, WOMAC total score of the right knee, WOMAC total score of the left knee, supplemental vitamins and minerals frequency, and antioxidant combination multivitamins frequency) that can be utilized in clinical practice to determine the risk of knee OA progression in individual patients. We also developed a prognostic model to estimate the risk for total knee replacement and provide suggestions for modifiable variables to improve long-term knee health. This combination of unsupervised and supervised data-driven tools provides a framework to identify knee OA phenotype in a clinical scenario and personalize treatment strategies.

## Introduction

Osteoarthritis (OA) is the most common form of joint disease and a major cause of pain and disability and is a heterogenous disease in which aging, obesity, trauma, and genetic factors are implicated as drivers of pathogenesis^1^. OA affects 9.6% of men and 18% of women over 60 years of age^2^ and 250 million people worldwide^3^. The United States Food and Drug Administration (FDA), Centers for Disease Control (CDC), and National Institutes of Health (NIH) all recognize the impact of OA and have guidelines and research agendas to reduce the prevalence and burden. This public health issue is projected to worsen as life expectancy increases and the US population skews towards older individuals^4^. Still, there are no disease-modifying OA drugs (DMOADs) approved by the FDA or European Medicines Agency^5^ and as a result, managing OA remains largely palliative.

One complicating factor is that OA phenotypes vary from patient to patient and there is likely no “one size fits all” treatment^6^. It may be that the failure of numerous phase II/III OA clinical trials, such as iNOS^7^, bisphosphonates^8^, and calcitonin^9, 10^, has been due to the inability to decipher the specific underlying drivers of OA at the individual patient level and therefore DMOADs are not delivered to the most suitable subgroups. Thus, identifying OA phenotypes is a critical task for the community. Machine learning (ML) is a computational tool that learns complex non-linear patterns between many variables without precise instructions^11 12, 13^. Classification ML models can identify novel, clinically significant features in patients^14, 15^. These methods have been used to determine disease phenotypes in many clinical populations^16^. Furthermore, predictive ML models have been used to determine disease risk factors, complications, and survival outcomes in clinical practice^17^. Our global hypothesis is that the heterogeneity in knee OA phenotypes can be organized with unsupervised learning and that supervised learning models can predict disease progression.

In the current study, we used unsupervised and supervised ML methods to identify knee OA phenotypes and predict disease progression in the open access Osteoarthritis Initiative (OAI) dataset (**Figure 1**). The OAI is a longitudinal, observational study of knee OA with 4,796 enrollees. It includes greater than 1,000 descriptive variables, including demographics, pain, exercise habits, diet and nutrition, socioeconomic status, medical history, radiographic evaluation, and psychological evaluation. We determined OA phenotypes by performing unsupervised learning on enrollment data (k-means clustering) and visualized relationships between phenotypes via dimensionality reduction. Then, we utilized data from multiple follow-up time points over 8 years to develop supervised learning models that predicted long-term disease progression, including the likelihood of total knee replacement (TKR).

**Figure 1.**
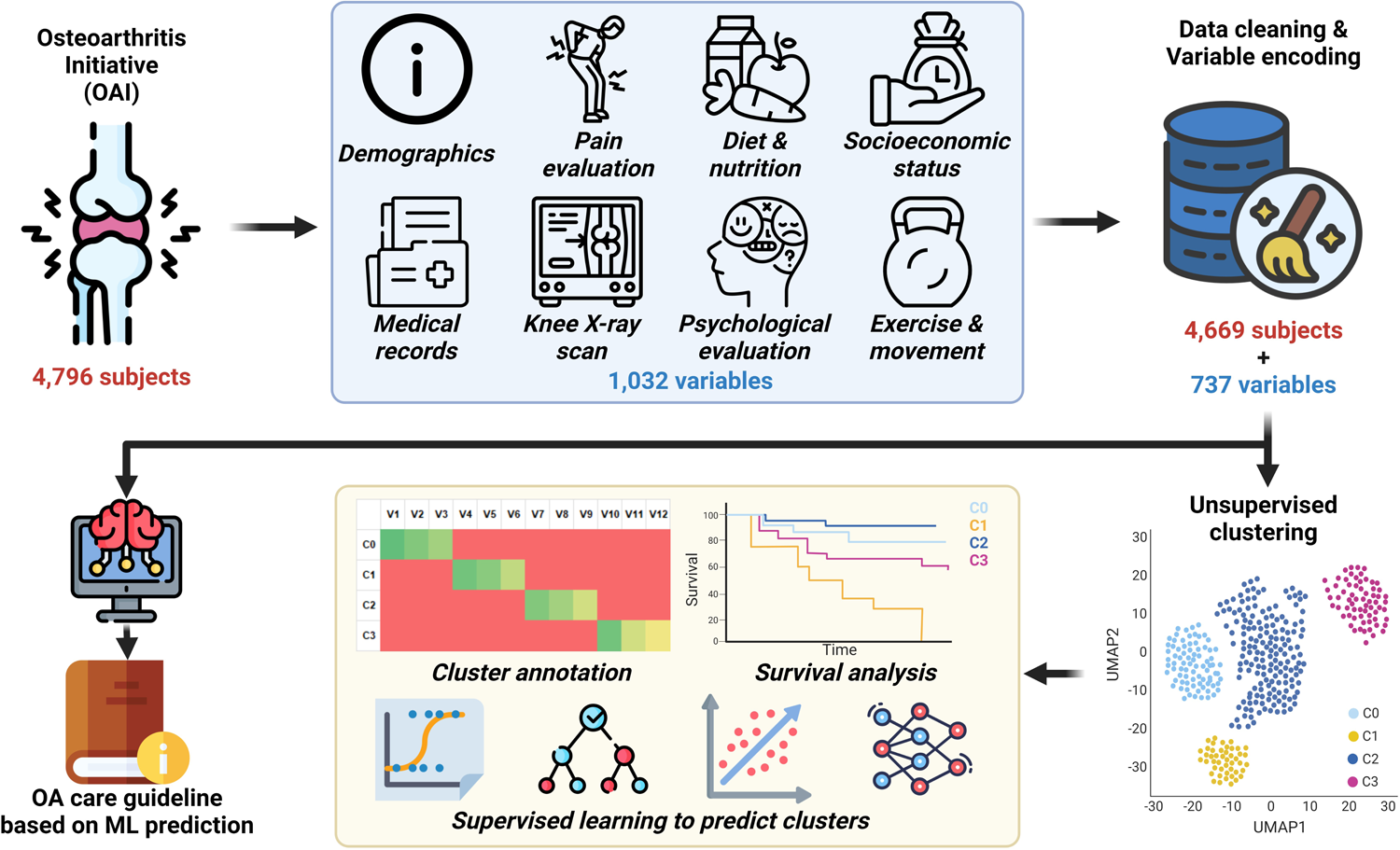
Overview of the experiment design. Osteoarthritis Initiative (OAI) data was organized and cleaned with 4,669 subjects (patients) and 737 variables. Unsupervised clustering was used to stratify the patients into four clusters. The detailed characteristics of each cluster were investigated with cluster annotation and survival analysis. A web-based clinical tool was developed to predict the cluster new patient could belong to with required information. Based on the most accurate WOMAC total score (WOMTS) prediction from an artificial intelligence model, and OA care guideline was also provided for translational usage.

## Materials and Methods

### Data extraction and cleaning

We included all 4,796 participants who enrolled in the OAI study with 1032 variables that were measured at enrollment (variables: **Data S1**). We performed a data cleaning procedure to remove individuals with incomplete data, remove variables that had missing values or low variance, and remove variables that were highly correlated. All data was processed in either Python or R as noted below.

First, we excluded 127 subjects with more than 595 variables (50% of total variables) whose value were missing (**Figure S1A**). Next, we generated a correlation matrix for each combination of numerical and categorical variables with the following calculations: 1) numerical vs. numerical: Pearson’s coefficient (pearsonr function from scipy.stats Python library, V1.10.1); 2) categorical vs. categorical: Cramers’ V (customized function based on Python); 3) numerical vs. categorical: R value from ordinary least squares liner regression (ols function from statsmodels.formula.api Python library, V0.13.5). We performed hierarchical clustering (Heatmap function from ComplexHeatmap R package, V2.14.0) to group variables in the correlation matrix and found that variables with missing values were grouped together. We screened different cutoffs (i.e., 25%, 50%, 80%) for the relative subject number of missing values (number of subjects with missing value relative to total subject number) and found that a 25% cutoff removed clustered variables with majority missing values (**Figure S1B, C, and D**) (**Data S1**). Therefore, we removed 295 variables among which more than 25% data points were missing (**Figure S1A**).

### Clustering and dimensionality reduction for identifying knee OA phenotypes

After data extraction and cleaning, we identified groups of similar individuals via unsupervised learning and performed dimensionality reduction for data visualization. First, we used the one-hot encoding method (get_dummies function from pandas Python library, V1.5.3) to convert the categorical variables to numerical variables. We replaced missing values using a k-Nearest Neighbors imputation (KNNImputer function from sklearn.impute Python library, V1.2.2) with 2 neighboring samples and uniform weights. Imputed data was scaled and normalized (StandardScaler function from sklearn.preprocessing Python library, V1.2.2) and principal component reduction was performed (PCA function from sklearn.decomposition Python library, V1.2.2). Based on the elbow method for variance thresholding (**Figure S2A**), the top 16 principal components were selected for dimensionality reduction (Uniform Manifold Approximation and Projection, UMAP, umap function from umap-learn Python library, V0.5.3) and K-Means clustering (KMeans function from sklearn.cluster Python library, V1.2.2). We calculated Silhouette scores (**Figure S2B**, silhouette_score function from sklearn.metrics Python library, V1.2.2) for 2 to 21 clusters and identified that 4 was the optimal cluster number.

We performed statistical comparisons to identify variables that differentiated each cluster. We used the Kruskal Wallis test for numerical variables (kruskal.test function from stats R package, V4.2.3) and Fisher’s exact test for categorical variables (fisher.test function from stats R package, V4.2.3). P-values for both numerical and categorical variables were adjusted by Benjamini & Hochberg method (**Data S2**, adjust_pvalue function from rstatix R package, V0.7.2). We identified the top 10 variables that differentiated each cluster based on the following criteria: numerical variables: maximum fold difference between means, categorical variables: Chi-square statistic (chisq.test function from stats R package, V4.2.3). Cluster annotations were determined by authors based on these cluster markers.

### Long-term outcomes across clusters and cohorts

For the four clusters identified in our study and for the three cohorts defined at OAI data collection, we performed Kaplan-Meier (KM) survival analysis using data from enrollment and each follow-up visit on the following 6 outcome variables: Kellgren-Lawrence (KL) grade, joint space width (minimum joint space width in the medial compartment), Western Ontario and McMaster Universities Osteoarthritis Index (WOMAC) disability score, WOMAC stiffness score, WOMAC pain score, WOMAC total score (WOMTS). We defined a survival event as the change of each outcome variable from the first visit to any follow-up visit above a defined threshold (KL grade Δ≥1; joint space width Δ≤-25%; all WOMAC scores Δ≥25%). We used exact enrollment and visit dates to account for variability in time between visits (**Data S3**). Once a progression event was identified, all following visits were discarded. We also extracted whether an individual received a total knee replacement (TKR) in either knee, where TKR was considered as the survival event. With such converted survival information, we built KM curves for all outcome variables for both knees (surv and survfit function from survival R package, V3.5.5). To quantify the hazard ratios for each cluster, we built Cox regression models (coxph function from survival R package, V3.5.5).

To further examine the prognostic values of our clusters, we implemented the same KM survival analysis on all four clusters within the progression cohort and incidence cohort separately. We built KM curves for all outcomes variables for both knees (surv and survfit function from survival R package, V3.5.5). To quantify the hazard ratios for each cluster within these two cohorts, we built Cox regression models (coxph function from survival R package, V3.5.5).

### Development of a clinical tool to predict cluster assignment via supervised learning

With well-defined clusters and survival outcomes by cluster, we developed a clinical tool that assigns individual patients to the appropriate cluster to determine their long-term knee health. To do so, we benchmarked common supervised learning models to predict cluster assignment. We evaluated logistic regression (LogisticRegression function from sklearn.linear_model Python library, V1.2.2; solver: newton-cg solver, maximum iterations: 1000), random forest (RandomForestClassifier function from sklearn.ensemble Python library, V1.2.2; trees: 100, entropy criterion), and support vector machine (SVC function from sklearn.svm Python library, V1.2.2; kernel: sigmoid, probability estimation enabled).

As above, we utilized numerical data and one-hot encoded categorical data as input data. We scaled each variable to its corresponding minimum and maximum range (MinMaxScaler function from sklearn.preprocessing Python library, V1.2.2). To determine the optimal number of input variables, we first ranked input variables based on the importance metrics calculated by fitting a random forest classifier (RandomForestClassifier function from sklearn.ensemble Python library, V1.2.2) to all the input variables with the cluster labels (**Data S4**) and then screened the input variable number from 2 to 50 for all ML models. To obtain robust accuracies of each input variable number, we utilized a random permutation cross-validator with 20 splits, and within each split, 90% samples were considered as training data while the left 10% were validation data (ShuffleSplit function from skleran.model_selection Python library, V1.2.2). As a multi-classification problem, we computed the accuracy classification score (accuracy_score function from sklearn.metrics Python library, V1.2.2) and area under the receiver operating characteristic curve (ROC AUC) using both one-vs-rest and one-vs-one approaches (roc_auc_score function from sklearn.metrics Python library, V1.2.2) (**Data S5**). We averaged the above metrics across all 20 test splits for each input variable number.

The most accurate model was exported and built on a web-based interface (www.predictoaphenotpe.org). With free registration, users will be able to fill in required information of the patient and the website will provide a prediction of the cluster (phenotype) this patient could belong to.

### Supervised learning for predicting WOMTS and identifying key predictor variables

We benchmarked common supervising learning models to predict WOMTS at 4 and 8 years from enrollment data. To identify effective predictor variables, we computed the correlations between input variables and WOMTS across all yearly visits for both knees. We computed Spearman’s correlation coefficients (spearmanr function from scipy.stats.stats Python library, V1.10.1) or R from ordinary least squares regression (ols function statsmodels.formula.api Python library, V0.13.5) to quantify the correlation between WOMTS and numerical or categorical variables (**Data S6**). We visualized the top 10 highly correlated variables based on their average correlation coefficients.

We directly predicted the WOMTS for both knees at 4^th^ and 8^th^ year visit. We evaluated linear regression (LinearRegression function from sklearn.linear_model Python library, V1.2.2), random forest (RandomForestRegressor function from sklearn.ensemble Python library, V1.2.2; trees: 10, 20, 40, 60, 80, 100), support vector machine (SVR function from sklearn.svm Python library, V1.2.2; kernels: linear, polynomial, rbf, sigmoid; regularization: 100, kernel coefficient: reciprocal of variable number), and an artificial neural network (ANN). We followed the same scaling, input variable selection, and cross-validation procedures used in predicting clusters. As WOMTS is a continuous variable, all ML models were regression models and used to compare the measured and predicted WOMTS we calculated root mean square error (RMSE, mean_squared_error function from sklearn.metrics Python library, V1.2.2) and Pearson’s correlation coefficient (PCC, pearsonr function from scipy.stats Python library, V1.10.1) as accuracy metrics (**Data S7**). Average accuracy metrics across all cross-validation tests were calculated to select the optimal input variable number.

For the ANN, we built a sequential model (Sequential function from keras.moedls Python library, V2.11.0) with one input layer, adaptive hidden layers, and one output layer (Dense function from keras.layers Python library, V2.11.0). The node number of the input layer was dependent on the number of input variables during the screening, and the output layer had one node to represent the WOMTS. The hidden layers were adaptively designed based on the number of input variables, where each hidden layer was 75% of its previous layer (including input layer). All activation functions were linear functions, Adam optimization with 0.001 as the learning rate (optimizer.Adam function from tensorflow.keras Python library, V2.11.0) was used to train the model, and mean squared error was taken as the loss function. We trained the model with 100 epochs and 10 as the batch size.

As the ANN achieved the most accurate and robust predictions, we utilized the ANN model to identify the most effective predictor variables using a customized random search algorithm. We firstly built the same sequential model (Sequential function from keras.models Python library, V2.11.0) with one input layer, adaptive hidden layers, and one output layer, adaptive hidden layers, and one output layer (Dense function from keras.layers Python library, V2.11.0) as the above ANN model. The node number of the input layer was 25 based on the screening results, and the output layer had one node to represent the WOMTS. Similarly, the hidden layers were adaptively designed based on the number of input variables, where hidden layer was 75% of its previous layer (including input layer). All activation functions were linear function, Adam optimization with 0.001 was the learning rate (optimizer. Adam function from tensorflow.keras Python library, V2.11.0) was used to train the model, and mean squared error was taken as the loss function. We trained the model with 100 epochs and 10 as the batch size. Here, we randomly selected 25 variables to train an ANN model based on the above design principles. To reduce the number of potential combinations, we only selected variables from the cluster markers identified from the unsupervised clustering (adjust p-value <0.05, **Data S2**). Within each test, we also used the same random perturbation cross-validator with the same parameters to obtain the accuracies. After 10,000 random selection tests, we ordered the test based on their average prediction accuracy and selected the top 10 to 1,000 most accurate tests to investigate the composition of their input variables. We quantified the popularity of each variable by computing the relative occurrences of each variable within the most accurate tests to the total 10,000 tests.

### Additional Statistics

Graphs and statistics were performed using R (v4.2.3), and Python (v3.9.16) as described. The Kruskal Wallis test, Fisher’s exact test, and log-rank test were implemented to compare numeric, categorical, and survival data across different phenotypes or cohorts. Pearson’s correlation coefficient, Spearman’s correlation coefficients, and Cramer’s V were calculated to quantify the associations. Accuracy, one-vs-one, and one-vs-rest AUC were calculated from multi-class prediction. Root-mean-square-error and correlation between prediction and measurements were calculated for regression. Experiment specific detailed statistical methods are described in corresponding figure legends and Methods sections. Calculated p values are displayed as *, p<0.05; **, p<0.01; ***p<0.001; ****, p<0.0001.

### Data and code availability

All scripts used in this publication are available in https://github.com/weihuaguo/cluster_oai. All other data are available in the main text or the supplementary materials.

## Results

### Unsupervised learning identified four knee OA phenotypes in OAI

We identified four knee OA phenotypes by unsupervised learning: a group with low supplemental and dietary vitamin intake (‘Low Vitamin’), a group with poor knee health (‘Poor Knee’), a group with intermediate knee health (‘Intermediate Knee’), and a group with good knee health (‘Good Knee’) (**Figure 2A**, **Table 1**). These names are based on the most significant and abundant variables between the groups (**Figure 2B&C, Figure S3A&B**). Specifically, the Low Vitamin group was characterized by low frequency of vitamin supplementation and low percentage of vitamins obtained from daily food intake, despite demonstrating good knee health and daily function. The Poor Knee group was characterized by poor knee health, in addition to low quality of life, poor general health, and poor daily function. The Intermediate Knee group exhibited relatively poor knee health, intermediate quality of life, and intermediate daily function. Lastly, the Good Knee group demonstrated good knee health, along with good quality of life, good general health, good mental health, and good daily function, (**Figure 2D-I**).

**Figure 2.**
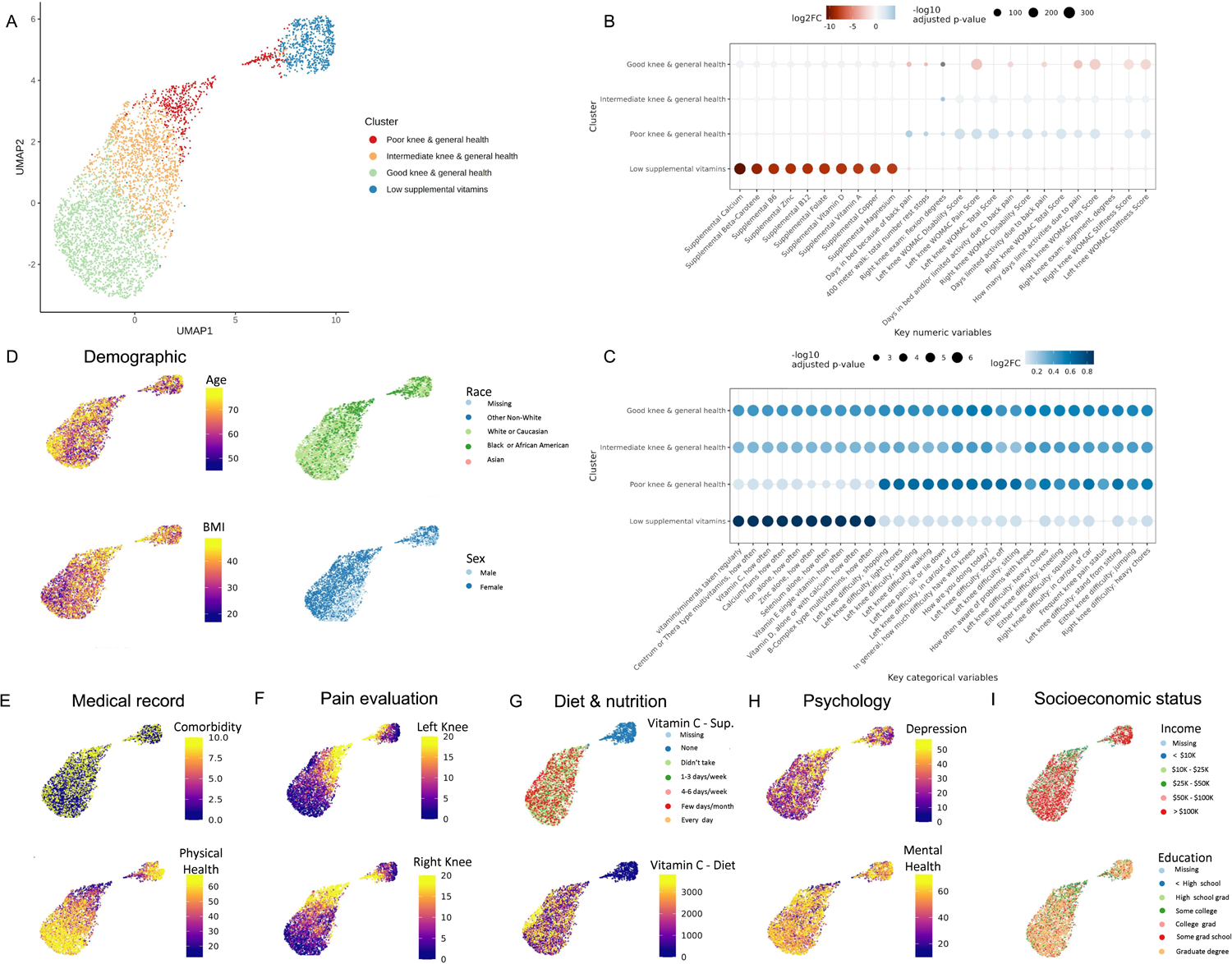
Cluster characteristics of OAI. (A) Four clusters on UMAP. (B) Top 10 numeric variables of each cluster. Kruskal-Wallis test was used to determine the statistics between the cluster of interests and all the other clusters together. Benjamini & Hochberg method was used to adjust the p-value. The numerical variables with adjusted p-values <0.05 were ranked by the log2 fold changes (log2FC) to select the top 10 of each cluster. (C) Top 10 categorical variables of each cluster. Fisher’s exact test was used to determine the statistics between the cluster of interests and all the other clusters together. Benjamini & Hochberg method was used to adjust the p-value. The categorical variables with adjusted p-values <0.05 were ranked by the Pearson’s chi-squared statistics to select the top 10 of each cluster. (D)∼(I) Key variables categorized into demographic (V00AGE, age; P02RACE, race; P02SEX, gender; P01BMI, BMI at baseline), medical record (V00COMRB, Charlson Comorbidity Index; V00HSPSS, Short Form 12 Physical Summary Score), pain evaluation (V00WOMKPL/R, WOMAC pain score of left/right knee), diet & nutrition (V00VITCCV, Vitamin C single vitamin, how often taken in past 12 months; V00SUPVITC, average daily Vitamin C supplement, mg), psychological evaluation (V00CESD, Center for Epidemiology Studies Depression Index; V00HSMSS, Short Form 12, Mental Summary Score), socioeconomic status (V00INCOME, annual personal income; V00EDCV, education level) on UMAP.

**Table 1.**

| Table 1 | Poor knee &<br>general health<br>(N=531) | Intermediate knee<br>& general health<br>(N=1359) | Good knee &<br>general health<br>(N=2096) | Low supplemental<br>vitamins<br>(N=683) | Overall<br>(N=4669) |
| --- | --- | --- | --- | --- | --- |
| Age |  |  |  |  |  |
| Mean (SD) | 60.1 (8.91) | 61.7 (9.11) | 62.1 (9.03) | 58.9 (9.42) | 61.3 (9.17) |
| Median [Min, Max] | 59.0 [45.0, 79.0] | 62.0 [45.0, 79.0] | 62.0 [45.0, 79.0] | 56.0 [45.0, 79.0] | 61.0 [45.0, 79.0] |
| Race |  |  |  |  |  |
| 0: Other Non-white | 18 (3.4%) | 27 (2.0%) | 26 (1.2%) | 10 (1.5%) | 81 (1.7%) |
| 1: White or Caucasian | 215 (40.5%) | 1090 (80.2%) | 1866 (89.0%) | 520 (76.1%) | 3691 (79.1%) |
| 2: Black or African<br>American | 295 (55.6%) | 228 (16.8%) | 184 (8.8%) | 145 (21.2%) | 852 (18.2%) |
| 3: Asian | 3 (0.6%) | 11 (0.8%) | 20 (1.0%) | 7 (1.0%) | 41 (0.9%) |
| .: Missing/Incomplete | 0 (0%) | 3 (0.2%) | 0 (0%) | 1 (0.1%) | 4 (0.1%) |
| BMI |  |  |  |  |  |
| Mean (SD) | 31.7 (5.36) | 29.2 (4.56) | 27.5 (4.48) | 29.0 (4.57) | 28.7 (4.81) |
| Median [Min, Max] | 31.3 [18.8, 48.7] | 29.0 [17.2, 47.7] | 27.0 [16.9, 46.8] | 28.6 [17.8, 44.6] | 28.3 [16.9, 48.7] |
| Missing | 2 (0.4%) | 2 (0.1%) | 0 (0%) | 0 (0%) | 4 (0.1%) |
| Education |  |  |  |  |  |
| 0: Less than high school<br>graduate | 79 (14.9%) | 27 (2.0%) | 37 (1.8%) | 24 (3.5%) | 167 (3.6%) |
| 1: High school graduate | 123 (23.2%) | 188 (13.8%) | 204 (9.7%) | 89 (13.0%) | 604 (12.9%) |
| 2: Some college | 196 (36.9%) | 369 (27.2%) | 420 (20.0%) | 147 (21.5%) | 1132 (24.2%) |
| 3: College graduate | 49 (9.2%) | 271 (19.9%) | 508 (24.2%) | 150 (22.0%) | 978 (20.9%) |
| 4: Some graduate school | 33 (6.2%) | 116 (8.5%) | 195 (9.3%) | 48 (7.0%) | 392 (8.4%) |
| .: M Missing/Incomplete | 2 (0.4%) | 1 (0.1%) | 0 (0%) | 1 (0.1%) | 4 (0.1%) |
| Income |  |  |  |  |  |
| 1: Less than \$10K | 80 (15.1%) | 34 (2.5%) | 29 (1.4%) | 17 (2.5%) | 160 (3.4%) |
| 2: \$10K to < \$25K | 109 (20.5%) | 129 (9.5%) | 154 (7.3%) | 59 (8.6%) | 451 (9.7%) |
| 3: \$25K to < \$50K | 150 (28.2%) | 362 (26.6%) | 468 (22.3%) | 145 (21.2%) | 1125 (24.1%) |
| 4: \$50K to < \$100K | 100 (18.8%) | 464 (34.1%) | 786 (37.5%) | 227 (33.2%) | 1577 (33.8%) |
| 5: \$100K or greater | 31 (5.8%) | 291 (21.4%) | 517 (24.7%) | 199 (29.1%) | 1038 (22.2%) |
| .: Missing/Incomplete | 61 (11.5%) | 79 (5.8%) | 142 (6.8%) | 36 (5.3%) | 318 (6.8%) |
| Employment |  |  |  |  |  |
| 1: Works for pay | 277 (52.2%) | 795 (58.5%) | 1285 (61.3%) | 483 (70.7%) | 2840 (60.8%) |
| 2: Unpaid work for family business | 4 (0.8%) | 17 (1.3%) | 25 (1.2%) | 8 (1.2%) | 54 (1.2%) |
| 3: Not working in part due to health | 117 (22.0%) | 58 (4.3%) | 46 (2.2%) | 21 (3.1%) | 242 (5.2%) |
| 4: Not working other reasons | 125 (23.5%) | 480 (35.3%) | 736 (35.1%) | 170 (24.9%) | 1511 (32.4%) |
| .: Missing/Incomplete | 8 (1.5%) | 9 (0.7%) | 4 (0.2%) | 1 (0.1%) |  |

### Knee OA phenotypes are associated with disease progression

Survival analysis using KL grade (**Figure 3A**), joint space width (**Figure 3B**), WOMAC total score (**Figure 3C**), WOMAC pain score (**Figure 3D**), WOMAC stiffness score (**Figure 3E**), and WOMAC function score (**Figure 3F**) showed that the Good Knee group in both right and left knees. KL grade matched these trends for both knees; however, joint space width matched this trend in the right knee (p=0.00027) but not the left knee (p=0.44). By directly comparing these outcomes through all visits, we found that Good Knee group always had the lowest KL grade (**Figure S4A**), highest joint space width (**Figure S4B**), lowest WOMAC total score (**Figure S4C**), lowest WOMAC pain score (**Figure S4D**), lowest WOMAC stiffness score (**Figure S4E**), and lowest WOMAC function score (**Figure S4F**) on average. More importantly, survival analysis with total knee replacement outcome showed that the Good Knee group had the highest survival probability (**Figure 3G**).

**Figure 3.**
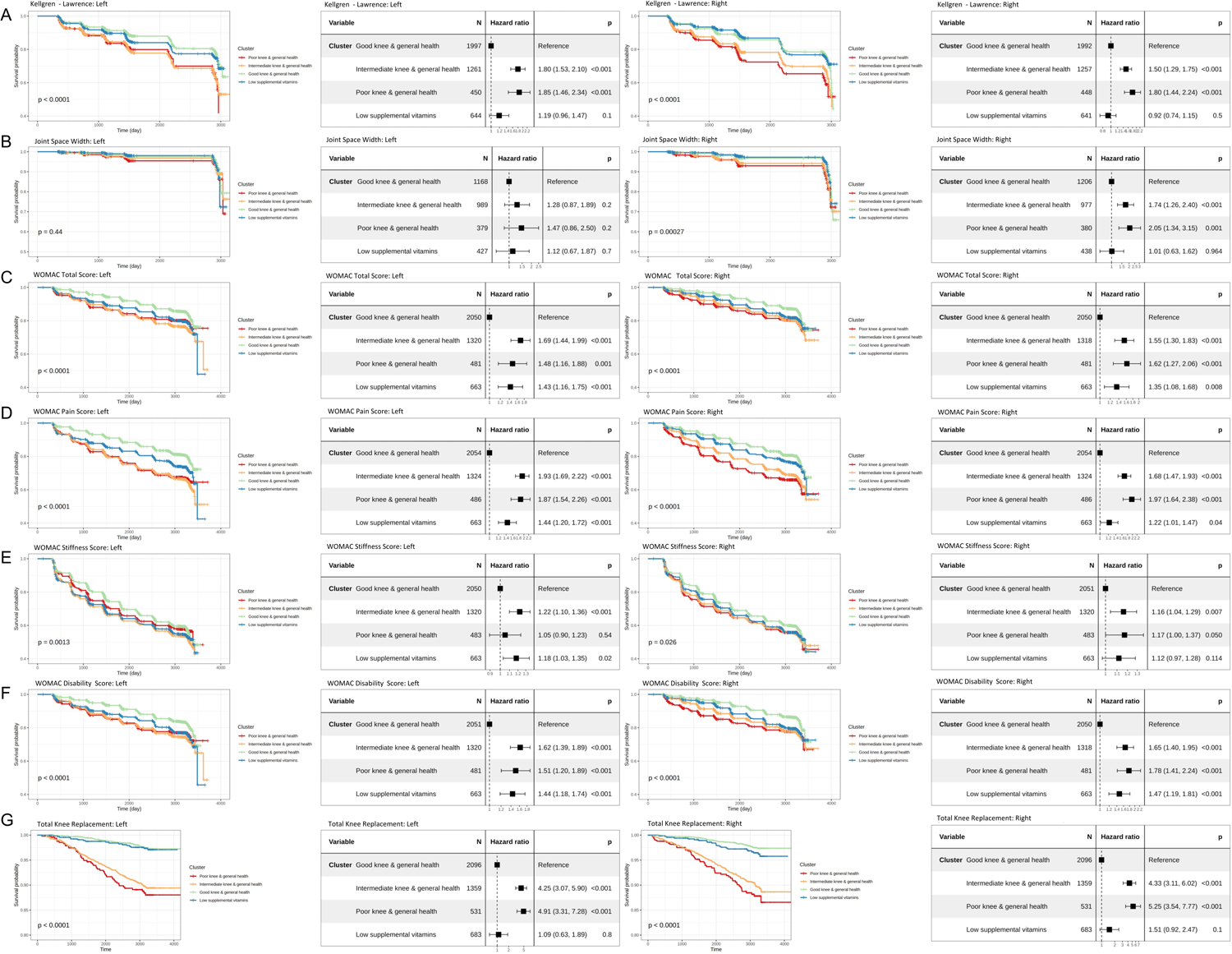
Prognostic values of 4 OA phenotypes. Kaplan-Meier plots (first and third from left) and forest plots (second and fourth from left) considering good knee health cluster as reference of KL grade (A), joint space width (B), WOMAC total score (C),WOMAC pain score (D), WOMAC stiffness score (E), WOMAC function score (F), and total knee replacement (G) were shown in a table format for both left (left two columns) and right (right two columns) knees. Log-rank p-value was shown in the KM plots. A univariant cox regression model for each outcome variable and each knee was built with the Good Knee group as the reference group and visualized in the forest plots.

Since the OAI has defined sub-cohorts (progression, incidence, and non-exposed control group)^18^, we first examined the composition of these sub-cohorts within our knee OA phenotypes (**Figure 4A**). We found that more than 85% of Good Knee subjects were from the incidence cohort, more than 60% of Poor Knee subjects were from the progression cohort, and more than 70% of Low Vitamin subjects were from incidence cohort. Since disease progression in these sub-cohorts were clinically well-defined, we tested our definitions of disease progression and survival by examining whether our cluster-based survival analysis results using patient WOMAC total scores (**Figure S5A**), KL grade (**Figure S5B**) joint space width (**Figure S5C**), and TKR (**Figure S5D**). As expected, our definition of disease progression and survival analysis comprehensively captured the disease progression based on the pre-defined sub-cohorts (i.e., non-exposed control group was the least progressed and progression cohort was the most progressed). Additionally, we also investigated the prognostic values of our knee OA phenotypes within incidence and progression sub-cohorts. The results showed that our knee OA phenotypes remained partially significant in patient WOMAC total scores (**Figure 4B**), KL grade (**Figure 4C**) and joint space width (**Figure 4D**), and TKR (**Figure 4E**) within the incidence and progression cohort. Except for joint space width, our four OA phenotypes tended to be associated with all the other clinical outcomes (p<0.10).

**Figure 4.**
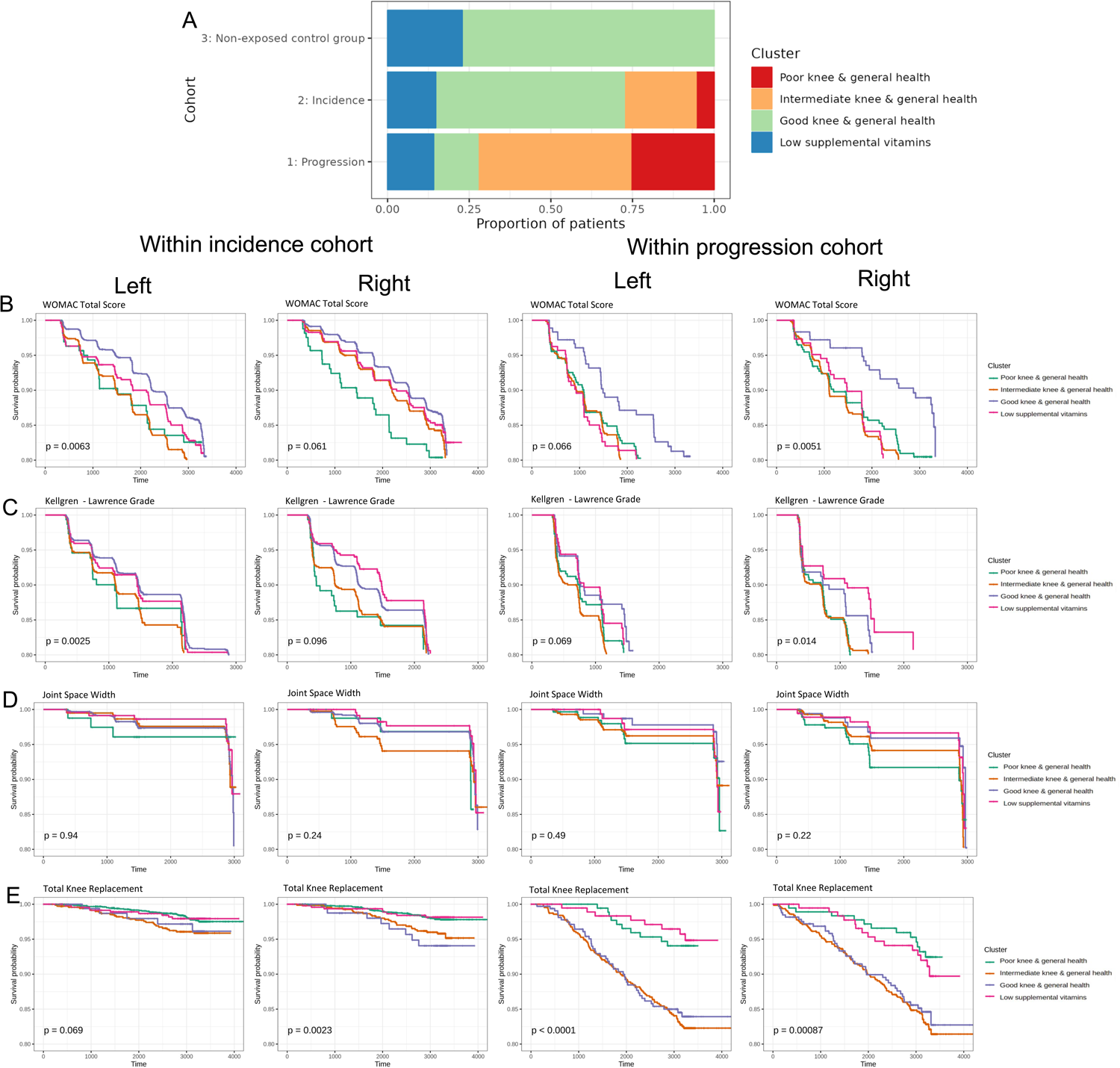
Prognostic values of four OA phenotypes within baseline cohorts. (A) Relative distribution of 4 OA phenotypes within each baseline cohort. Kaplan-Meier plots for WOMAC total score (B), KL grades (C), joint space width (D), total knee replacement (E) were shown in a table format for both left (left first and third columns) and right (left second and fourth columns) knees within incidence cohort (left two columns) and progression cohort (right two columns). Log-rank p-value was shown in the KM plots.

### Supervised learning accurately predicts cluster assignment

To accurately predict knee OA phenotypes, we benchmarked commonly used supervised learning models (four major types, i.e., logistic regression, random forest with six different tree numbers, supporting vector classifier with four different kernels). Generally, all models reached accuracy around 90%, above 0.975 AUC for ROC in both one-versus-one and one-versus-rest analyses (**Figure 5A**). Furthermore, we found that a minimum of five variables were necessary to achieve optimal predictive accuracy, namely: WOMAC disability score of the right knee, WOMAC total score of the right knee, WOMAC total score of the left knee, supplemental vitamins and minerals frequency, and antioxidant combination multivitamins frequency (**Figure 5B**).

**Figure 5.**
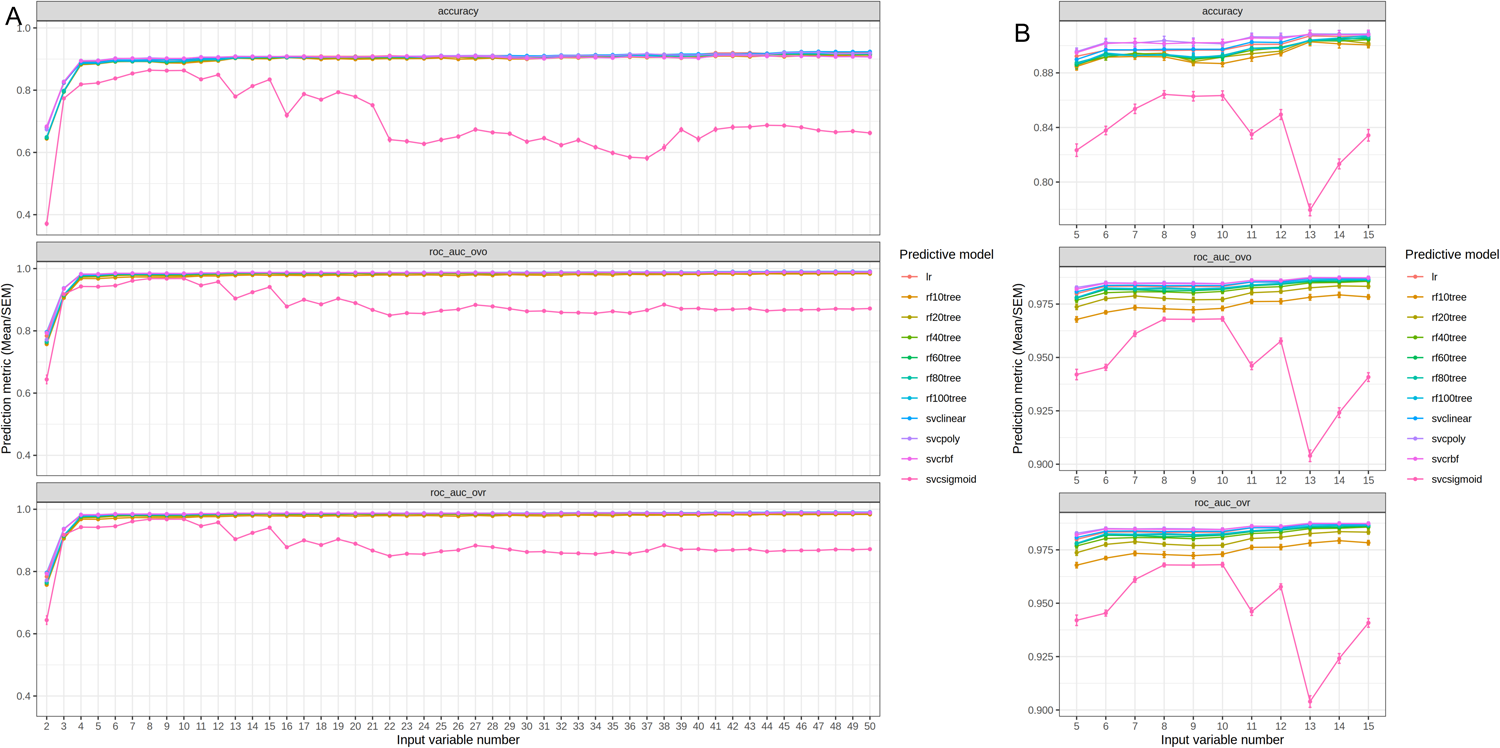
Prediction accuracies of cluster labels. A) Screening the optimal number of input variables (from 2 to 50) with different machine learning models (lr=linear regression, logistic regression model; rf10/20/40/60/80/100tree = random forest model with 10/20/40/60/80/100 trees; svclinear/poly/rbf/sigmoid = supporting vector classifier with linear/polynomial/radial basis function/sigmoid kernels). As a multi-class prediction problem, three accuracy metrics were used, i.e., accuracy (relative correct prediction numbers), roc_auc_ovo (area under curve of receiver operating characteristic curve, one vs one), and roc_auc_ovr (area under cuve of receiver operating characteristic curve, one vs rest). B) Detailed screening of the optimal number of input variables (from 5 to 15). The dot represents the mean of corresponding metric and the error bar represents the standard error of the mean from the cross-validation.

### WOMAC total score predictive modeling

Since WOMAC total score for both right and left knee is among the top variables for constructing accurate group prediction model, we first used univariate analysis to identify predictors at the screen phase (baseline) for the WOMAC total score of each visit (**Figure 6A**). The results showed that variables that were positively correlated were baseline right knee functional scores (difficulty in bathtub, standing, bending, car, shopping), baseline right knee WOMAC pain and disability scores, and baseline right and left knee WOMAC total scores. Variables that were negatively correlated include comorbidities and Knee Injury and Osteoarthritis Outcome Score (KOOS) scores (left and right knee KOOS pain, right knee KOOS quality of life, left and right knee KOOS symptom score).

**Figure 6.**
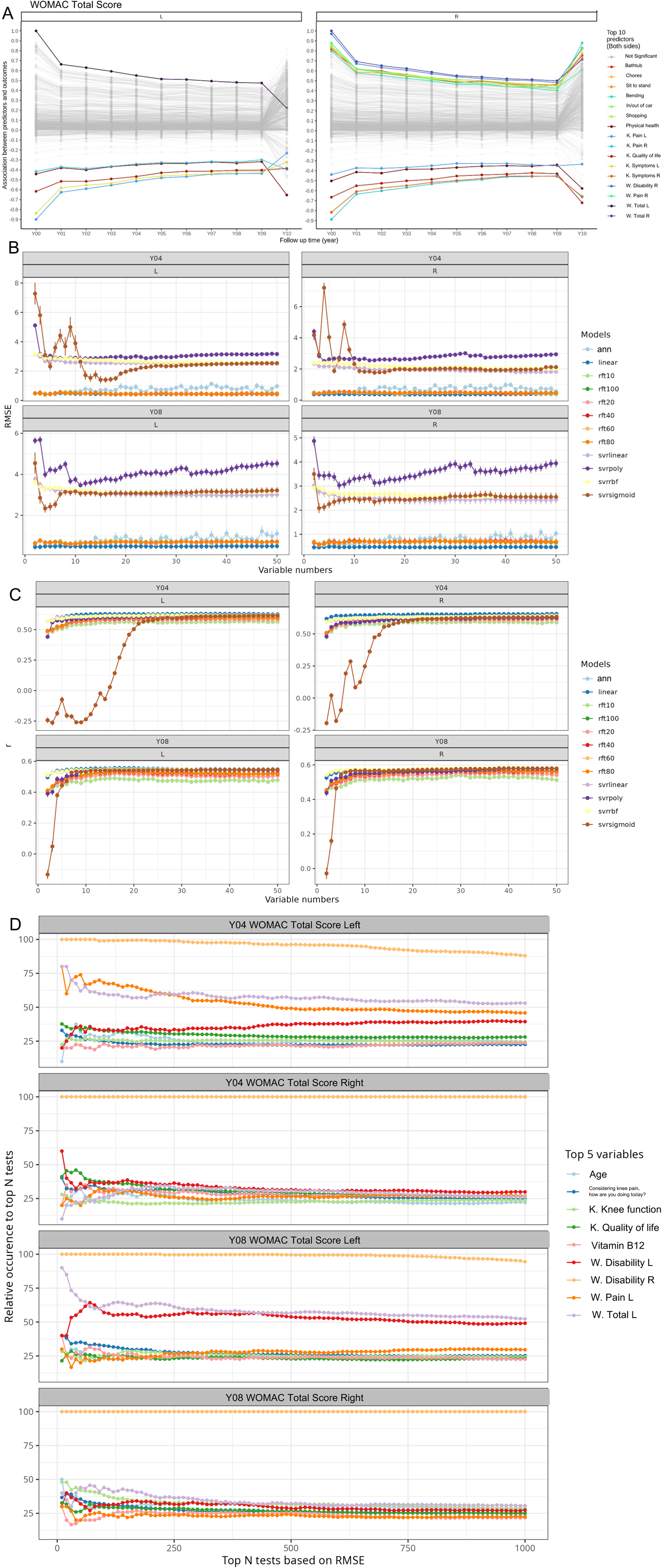
Prediction accuracy of WOMAC total score at fourth and eighth year. A) Correlation coefficients between WOMTS of all visiting years and baseline variables. The top 10 baseline variables were colored based on the average correlation coefficients crossing all the visiting years. W. = WOMAC, K.=KOOS. B) Screening the optimal number of input variables (from 2 to 50) with different machine learning models (linear, linear regression model; rft10/20/40/60/80/100 = random forest regressor with 20/40/60/80/100 trees; svrlinear/poly/rbf/sigmoid = supporting vector regressor with linear/polynomial/rbf/sigmoid kernels, ann = artificial neural network). As a regression problem, two accuracy metrics were used, i.e., RMSE (root mean squared error) and r (correlation coefficient between prediction and measurements). D) Top 5 variables with highest occurrences from the top 10000 most accurate prediction tests. We randomly selected 25 input variables from the cluster markers and used these variables to train an ANN model with the same settings with cross-validation. The above procedure was repeated 10,000 times. The top 1000 most accurate tests were extracted and the relative occurrence of each variable to these 1000 tests was calculated. The top 5 with highest relative occurrences for WOMTS of both left and right knees at fourth and eighth year were selected to visualize here. The dot represents the relative occurrences. W. = WOMAC, K.=KOOS

We then developed ML prediction-based multivariate analysis to identify a set of key variables related to WOMAC total scores (details in Methods). The principle of this analysis is that the input variables, which are necessary to accurately predict WOMAC total scores through ML models, are key variables. Based on this principle, we first benchmarked 12 multivariate supervised ML models on their accuracies in predicting WOMAC total scores for either knee. We found that the ML model built by ANN, linear regression, and an 80-tree random forest showed the best predictive accuracy reflected by lower RMSE (**Figure 6B**) and higher PCC between measurements and predictions (**Figure 6C**). Because ANN has the best robustness^19^, we utilize ANN as the ML model and randomly selected 25 input variables to train the ANN model and evaluate the corresponding prediction accuracy. With 10,000 random selections, we analyzed the relative occurrences of the input variables within most accurate predictions. Based on this analysis, we identified the top 5 variables with highest average occurrences from top 10 to 1,000 most accurate predictions of WOMAC total score at both 4-year and 8-year follow-ups. The results showed that variables had the greatest relative occurrence were age, iron supplement, knee difficulty – kneeling, difficulty with knees, B12 supplement, left knee WOMAC disability score, left knee WOMAC pain score, and left knee WOMAC total score. Among them, the baseline right knee WOMAC disability score had the greatest relative occurrence (**Figure 6D**).

## Discussion

OA is a heterogeneous disease and modern multivariate solution is likely necessary to identify disease phenotypes and progression patterns. In this study we identified four distinct knee OA phenotypes using unsupervised learning in the 4,796 participants of the Osteoarthritis Initiative. Phenotypes were primarily determined by nutrition and disability, stiffness, and pain (knee and back) scores and were strongly related to disease fate. In addition, we established a phenotyping tool from 5 variables that can be utilized in clinical practice to determine the risk of knee OA progression in individual patients. We also developed a prognostic model that can predict the risk of total knee replacement and provide suggestions for modifiable variables to improve long-term knee health.

We utilized all available subjects and variables from 10 years of follow-up data in the OAI. Our results show four distinct phenotypes that can be determined by simple questionaries related to general health, knee health, nutrition, and psychological evaluation. The groups included a group with a hallmark of low supplemental and dietary vitamin intake (‘Low vitamin’), a group with hallmarks of poor knee health (‘Poor Knee’), a group with hallmarks of intermediate knee health (‘Intermediate Knee’), and a group with hallmarks of good knee health (‘Good Knee’) (**Figure 2A**, **Table 1**). The names of these groups are based on the most statistically significant and prevalent variables between the groups (**Figure 2B&C**). Among them, the top variables were related to the frequency of vitamins/minerals intake, the amount of the supplemental Calcium, Beta-Carotene, Zinc, vitamin B6, B12, and D, WOMAC sub-scores, and WOMAC total score. Previously, other studies have tried to identify knee OA phenotypes. For example, by using biochemical markers data from IMI-APPROACH cohort, Angelini et al.^20^ found that OA patients could be divided into three phenotypes: low tissue turnover, structural damage, and systemic inflammation. In addition, by using RNA sequencing data from knee OA patients tissue (cartilage, subchondral bone, and synovium) Yuan et al.^21^ showed that OA patients could be divided into four subtypes: metabolic disorder subtype, collagen metabolic disorder subtype, activated sensory neuron subtypes, and inflammation subtype. In this work, we present a concise and clinically applicable OA phenotyping method that does not require intra-articular procedures, bloodwork, or sequencing that may be susceptible to error from environmental factors^22^.

Survival analysis revealed that the phenotypes defined by unsupervised learning were associated with long-term knee symptom, structure, and clinical outcomes (WOMAC total score, KL grade, TKR). More importantly, we developed phenotype prediction models and narrowed the necessary parameters down to 5 variables (WOMAC disability score, right knee; WOMAC total score, right knee; WOMAC total score, left knee; multivitamin frequency; antioxidant multivitamin frequency) which can be conveniently deployed in daily clinical scenarios. In the past, several studies tried to use ML methods to establish predictive models for TKR and achieved good accuracy^23–25^. However, there are limitations prohibiting these models from wide clinical use. Firstly, willingness to receive TKR is determined not only by medical related factors but also by others such as socioeconomic status and culture. Secondly, not all models served as a prognostic purpose. As OA is a chronic condition that is widespread, often ongoing, and frequently marked by episodes of exacerbation, long-term management of the disease is crucial for individualized treatment. Thus, the most important scientific question in this field is how to identify the appropriate patient for the correct treatment. Previously, Driban et al.^26^ utilized OAI data and found that 80% of people with end-stage knee OA did not have progressive radiographic severity, suggesting radiographic results alone are not an optimal variable for disease stage definition. In addition, Pierson et al.^27^ used an algorithmic approach and found out that radiologist-based X-ray interpretation could only explain 9% of unexplained racial disparities in pain, which makes determining the risk for TKR more difficult. We surmise that our approach which incorporates a holistic view of knee health is well-suited to a clinical setting. Interestingly, we identified a phenotype of Low Vitamin group with similar survival probability to the Good Knee group. The signature variables associated with the population from this group were the frequency of vitamin A and C intake. Antioxidant supplements such as vitamin A and C have long been advocated for the treatment of OA^28^. Although various approaches have been employed to tackle this issue, there is still a dearth of substantial evidence to support these treatments. In a systematic review, Canter et al.^29^ summarized 9 RCTs results and found that no convincing evidence to support vitamin A and C in OA treatment. Kraus et al. identified that Vitamin C can actually exacerbate OA in a guinea pig model^30^. In recent study, Qu et al.^31^ applied mendelian randomization to the data from UK Biobank and failed to find the causal association between vitamin A and OA. Our findings were supported by these data as a low supplemental vitamin intake did not worsen OA prognosis.

In the current study, other relevant factors like BMI, comorbidities, and depression statistically differentiated phenotypes in addition to signature variables mentioned above. BMI was one factor that contributed to the phenotypes and fate of the disease. This is consistent with the literature that suggests that BMI has long been considered as a risk factor for OA^32^. We also found that higher comorbidities were associated with worse knee OA phenotype and disease progression. Gustafsson et al.^33^ have shown that compared to matched references from the general population, knee OA patients were more commonly associated with one or more comorbidities, which was independent of socioeconomic status. We found the highest depression score and worst prognostic results in the Poor Knee group, which implies the importance of depression intervention in knee OA management. The association between mental health, especially depression, and knee OA has long been established. In an OAI sub-cohort, Rathbun et al.^34^ has reported the association between depression and faster disease progression and faster disease progression among individuals with radiographic knee OA. Additionally, in an older OA cohort, Parmelee et al.^35^ further confirmed depression as a moderator between OA pain and negative affect.

Our study has several strengths. First, our phenotyping models are parsimonious and do not rely on invasive or expensive genetic and biomarker outcomes. Secondly, all predictors can be collected when a patient seeks clinical care using validated questionnaires. Thirdly, the phenotyping we developed can predict long-term symptomatic and radiographic OA progression using modifiable predictors. Thus, it could be used to assist clinicians for clinical decisions to modify the risk factors and potentially lead to change of disease progression. Finally, our models were designed to not only address end-stage knee OA patients, but also individuals seeking clinical care due to recent knee pain, thus allowing for comprehensive disease cycle management.

Several limitations of our study are worth noting. First, although the OAI dataset used for our analysis enrolled a diverse patient group from sites across the USA, our findings need to be validated in independent populations. Secondly, in the current study, it was not possible to assess how using our phenotyping model as a decision aid would affect patient outcomes. However, we have built our phenotyping model into an online platform which can be openly accessed as validation step prior to its approval by regulatory bodies for clinical use.

## Conclusion

In summary, we identified four distinct knee OA phenotypes using unsupervised ML methods reflecting differences in knee symptoms and supplemental vitamin intake. Phenotypes were strongly associated with long-term disease fate. Supervised ML results confirmed that this phenotyping could be achieved with parsimonious, modifiable variables, and we propose this strategy could improve clinical decisions.

## Supporting information

Supplement Description

## Data Availability

All data produced in the present work are contained in the manuscript

## Acknowledgments

Dr. Zeyu Huang wishes to acknowledge funding from the National Natural Science Foundation of China (NSFC: 92049101; 81972097; 81702185) and Sichuan Science and Technology Programs (No. 2022YFH0101, 2018HH0071). Dr. John T. Martin wishes to acknowledge funding from the National Institute of Arthritis and Musculoskeletal and Skin Diseases (K99 AR077685, R00 AR077685).

## Author Contributions

John T. Martin, Zeyu Huang, and Weihua Guo designed the study. John T. Martin and Weihua Guo analyzed data. John T. Martin, Zeyu Huang, Mary A. Bucklin, and Weihua Guo interpreted the data. Zeyu Huang and Mary A. Bucklin drafted the manuscript. All authors critically revised the manuscript.

