## Supplement Description for "Disease progression and clinical outcomes in latent osteoarthritis phenotypes: Data from the Osteoarthritis Initiative"

### Supplementary Figures

**Supplementary Figure 1. Data cleaning and organization.** A) flowchart of sample and variable exclusion. B), C), and D) Heatmaps of quantitative correlation (left), corresponding p-value (middle), and number of available data pairs (right) with exclusion of the variables which have more than 25%, 50%, and 80% of missing subjects. The orders of rows and columns for all three heatmaps were obtained by hierarchical clustering of the correlation coefficients (left panel).

**Supplementary Figure 2. Parameter determination for clustering analysis.** A) Determining PC numbers based on the variance. B) Determining cluster numbers based on Silhouette scores.

**Supplementary Figure 3. Key features of each cluster.** A) Volcano plot for numeric variables B) Volcano plot for categorical variables. To determine the statistics, Kruskal Wallis test was used for numerical variables and Fisher's exact test was used for categorical variables. In Fisher's exact test, p-values were computed by Monte Carlo simulation with $10^7$ replicates. The p-values for both numerical and categorical variables were adjusted by Benjamini & Hochberg method (Data S3). The differences between the cluster of interest and all the other clusters were determined with log2 fold changes for numeric variables (A) and Chi-square statistic for categorical variables (B). Key cluster annotations were selected and labeled by authors.

**Supplementary Figure 4. Comparisons of outcome variables crossing four clusters throughout all the visits**. A) Stack barplot comparing the KL grades for both left (upper row) and right (lower row) knees crossing 4 clusters within each year visit. B) ~ F) Dotline plot comparing joint space width, WOMAC total score, WOMAC pain score, WOMAC stiffness score, and WOMAC function score for both left (upper) and right (lower) knees crossing 4 clusters within each year visit. The dot represents the average value, and the error bar represents the standard error of mean.

**Supplementary Figure 5. Survival analysis with baseline cohorts.** Kaplan-Meier plots for WOMAC total score (A), KL grades (B), joint space width (C), total knee replacement (D) were shown in a table format for both left (left column) and right (right column) knees. Log-rank p-value was shown in the KM plots.

### **Supplementary Data**

Supplementary Data 1. OAI variables utilized in analysis, including quality control designations.

Supplementary Data 2. All the markers of each cluster.

Supplementary Data 3. Alignment of visits with real yearly time.

Supplementary Data 4. Importance based on random forest classification for cluster labels.

Supplementary Data 5. Cluster prediction accuracies of cross-validations.

Supplementary Data 6. Correlation between WOMTS and other variables at all visits.

Supplementary Data 7. WOMTS prediction accuracies in 10,000 tests.
