## Supplementary figures and images for "Disease progression and clinical outcomes in latent osteoarthritis phenotypes: Data from the Osteoarthritis Initiative"

### Supplemental Figure 1

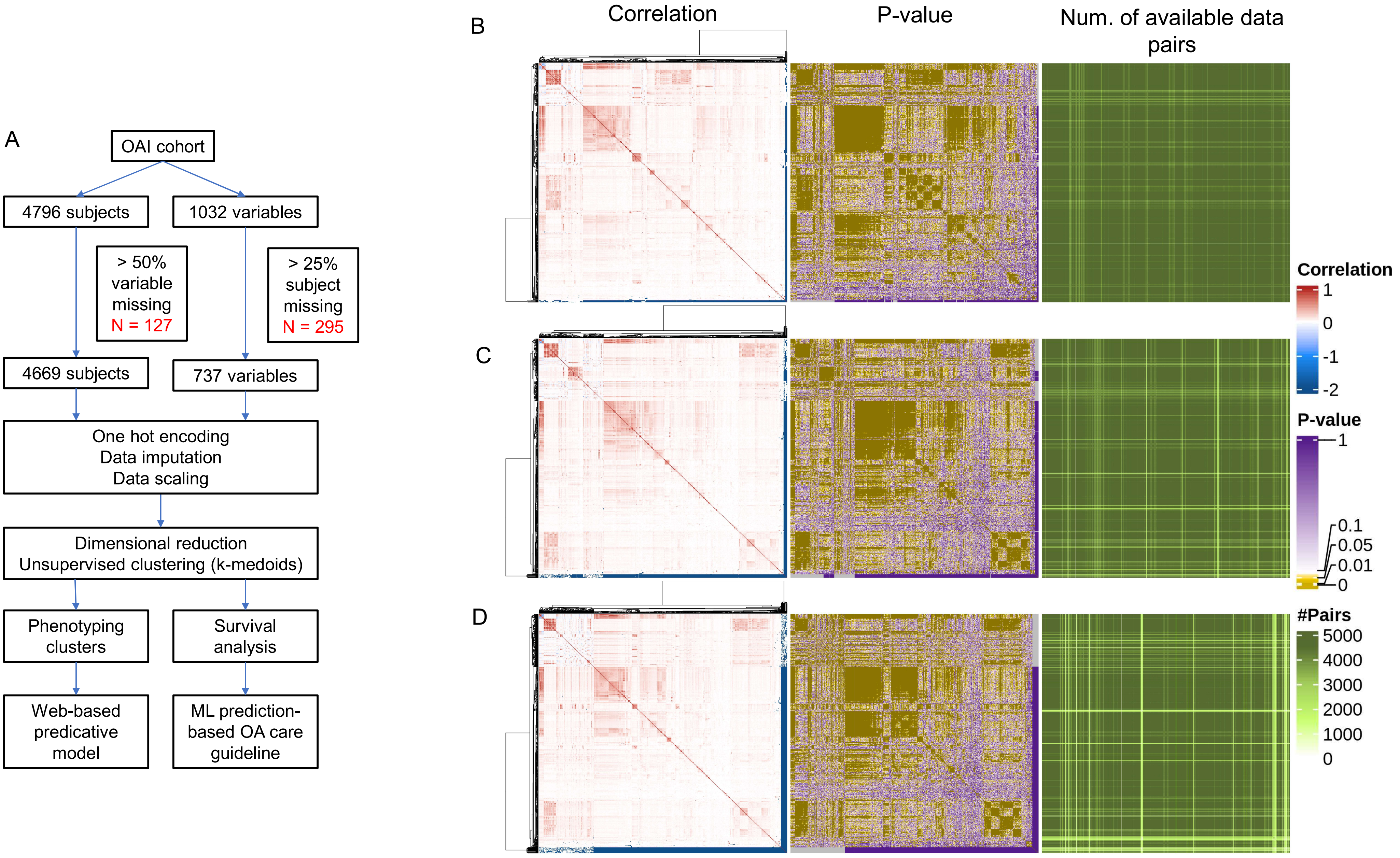

### Supplemental Figure 2

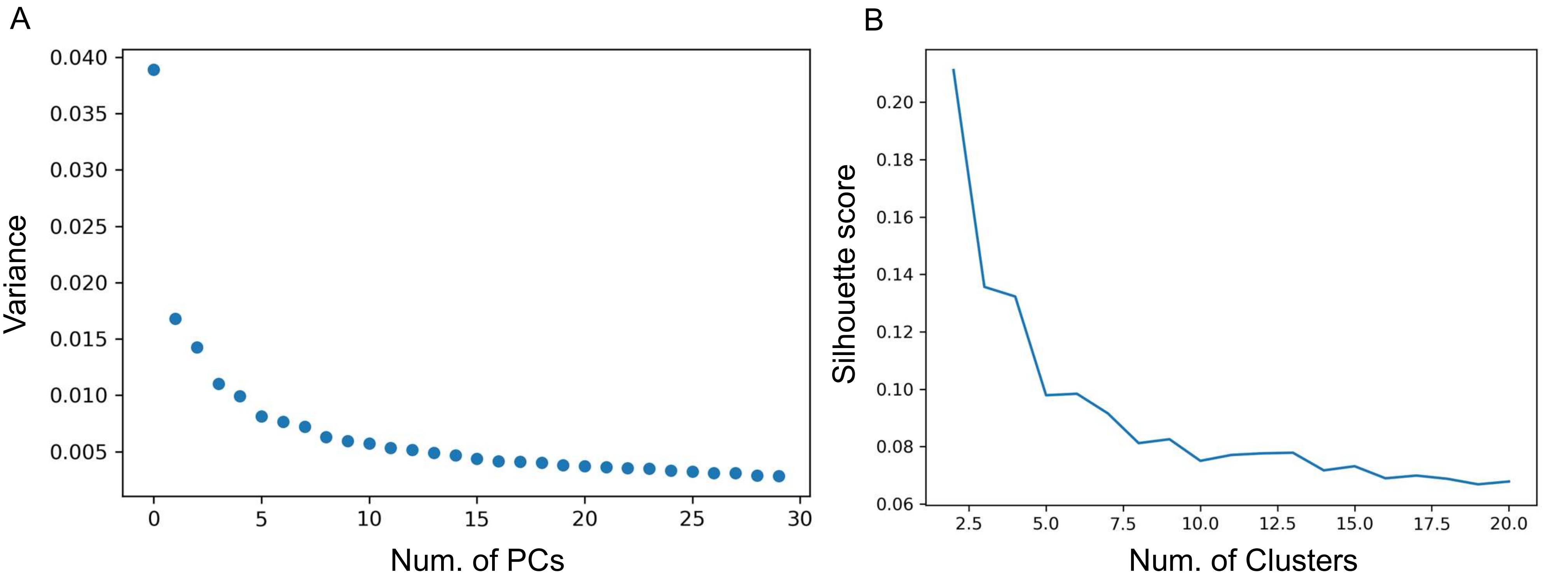

### Supplemental Figure 3

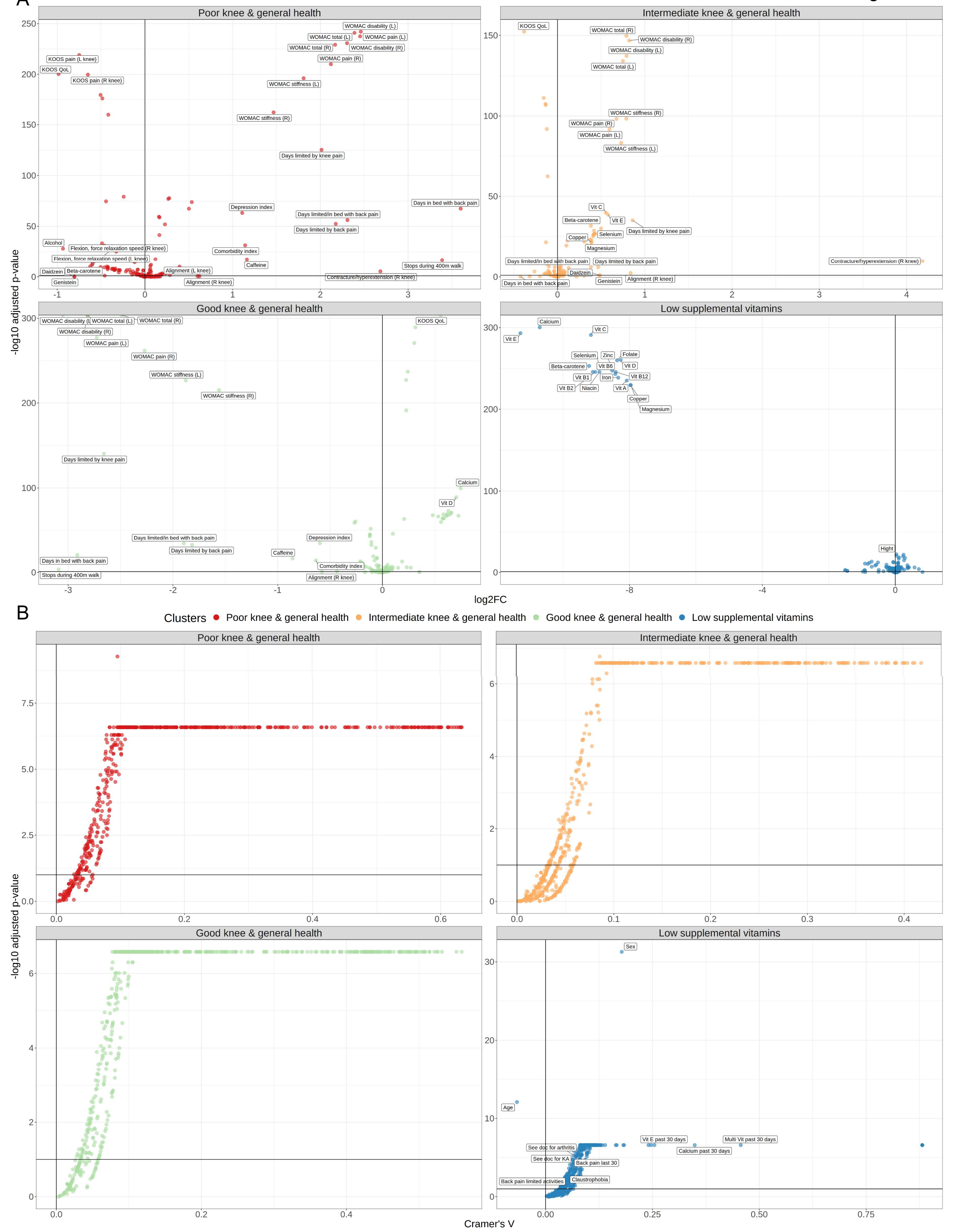

### Supplemental Figure 4

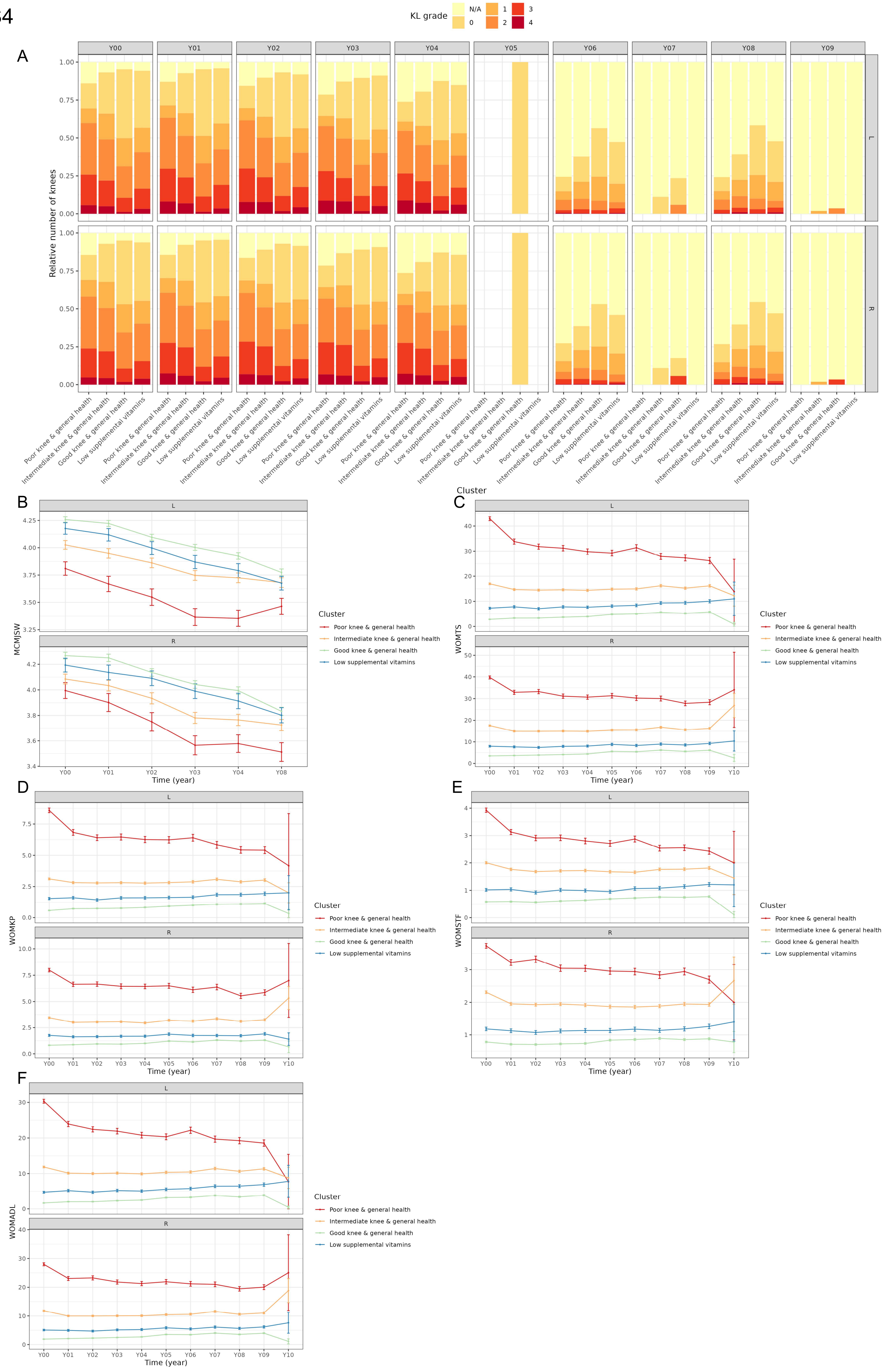

### Supplemental Figure 5

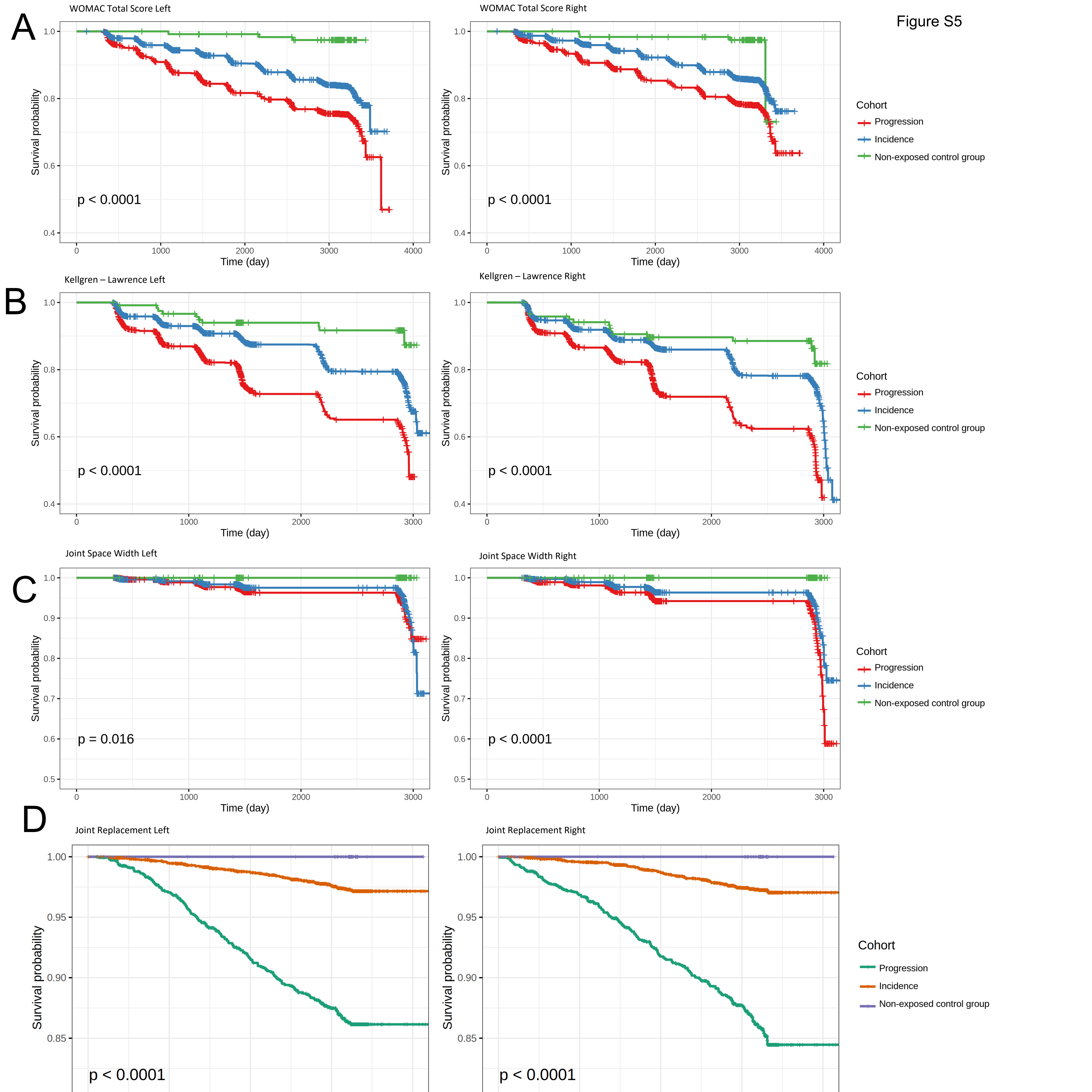
